# Greater working memory and speech perception scores in cochlear implant users predict better subjective quality of life and hearing

**DOI:** 10.1101/2022.09.07.22279622

**Authors:** Priyanka Prince, Joseph Chen, Trung Le, Vincent Lin, Andrew Dimitrijevic

## Abstract

A common concern in individuals with cochlear implants (CIs) is difficulty following conversations in noisy environments and social settings. The ability to accomplish these listening tasks relies on the individual’s working memory abilities and draws upon limited cognitive resources to accomplish successful listening. For some individuals, allocating too much, can result deficits in speech perception and in long term detriments of quality of life. For this study, 31 CI users and NH controls completed a series of online behavioural tests and quality of life surveys, in order to investigate the relationship between visual and auditory working memory, clinical and behavioural measures of speech perception and quality of life and hearing. Results showed NH individuals were superior on auditory working memory and survey outcomes. In CI users, recall performance on the three working memory span tests declined from visual reading span to auditory listening in quiet and then listening in noise and speech perception was predictably worse when presented with noise maskers. Bilateral users performed better on each task compared to unilateral/HA and unilateral only users and reported better survey outcomes. Correlation analysis revealed that memory recall and speech perception ability were significantly correlated with sections of CIQOL and SSQ surveys along with clinical speech perception scores in CI users. These results confirm that hearing condition can predict working memory and speech perception and that working memory ability and speech perception, in turn, predict quality of life. Importantly, we demonstrate that online testing can be used as a tool to assess hearing, cognition, and quality of life in CI users.

## 1. Introduction

In the population of cochlear implant users, speech perception, specifically in noise, is known to be variable across individuals and can depend on patient factors such as etiology, duration of deafness, age and device-related factors (Blamey et al., 1996; Holden et al., 2013; James et al., 2022; Lazard et al., 2012; Lazard & Giraud, 2017). The more significant factors are etiology and duration of deafness contributing ∼40% to the variability and device-related factors such as type of device and electrode position parameters, contributing ∼20% (James et al., 2022) however, quite a large amount is unaccounted for. Some studies have suggested that differences in individual cognitive processing are related to the variability of speech perception outcomes found in the CI population (Hast et al., 2015; Lenarz et al., 2012; Mahmoud & Ruckenstein, 2014).

This is evident in Skidmore et al. (2020), when including cognitive measures into a regression model to predict speech recognition. They found that at least 60% of speech recognition variance can be accounted for by four different demographic, sensory and cognitive measures and close to 80% accountable by twelve different demographic, sensory and cognitive measures (Skidmore et al., 2020). This might be attributed to a larger allocation of cognitive resources towards the encoding and the disambiguating of auditory stimuli as a result of spectrally degraded auditory signals from listening through a CI (Mattys et al., 2012; Ohlenforst et al., 2017; Rönnberg et al., 2010, 2016). Brain imaging studies corroborate this theory of increased use of the cognitive reserve in those with hearing loss, CI users and NH individuals presented with noise vocoded speech. Studies show a greater deployment of resources along with increased recruitment of temporal, parietal and frontal areas (Ala et al., 2020; Alain et al., 2018; Becker et al., 2013; Campbell & Sharma, 2013, 2014, 2020; Cartocci et al., 2018; Coez et al., 2014; Davis & Johnsrude, 2003; Decruy et al., 2020; Dimitrijevic et al., 2017; Glick & Sharma, 2017, 2020; Hervais-Adelman et al., 2012; Kessler et al., 2020; H.-J. Lee et al., 2007; Y.-S. Lee et al., 2016; Marsella et al., 2017; McMahon et al., 2016; Miles et al., 2017; Obleser & Weisz, 2012; Paul et al., 2021; Petersen et al., 2015) and modified connectivity (Bidelman et al., 2019; Campbell & Sharma, 2020; L. Chen et al., 2017; Davis & Johnsrude, 2007; Husain & Schmidt, 2014; Y. S. Lee et al., 2018; Murphy et al., 2020; Palva et al., 2010; Pessoa et al., 2002; Petrides & Pandya, 2002; Prince et al., 2021; Puschmann & Thiel, 2017; Schmidt et al., 2013; Sohoglu et al., 2012; Wild et al., 2012; Wolak et al., 2019; Zekveld et al., 2012) during perception, encoding, and retention of visual, auditory and verbal stimuli.

The effects of an increased recruitment of cognitive and working memory resources towards speech can lead to prolonged listening effort and an increased exhaustion of their cognitive reserves (Chang et al., 2004; Gates et al., 2011; Kathleen Pichora-Fuller et al., 1995; Lee et al., 2004; Lin et al., 2012; Lin et al., 2008; Rönnberg et al., 2013; Tun et al., 2009; Wingfield, 2016; Wu et al., 2009) which may, in turn, result in fatigue, social isolation, depression and overtime, cognitive decline (Akeroyd, 2008; Fritze et al., 2016; Fulton et al., 2015; Gallacher et al., 2012; Golub, 2017; Gurgel et al., 2014; F. R. Lin et al., 2013, 2014; Loughrey et al., 2018; Strawbridge et al., 2000; Wingfield, 2016). Various studies have found that hearing loss and cognitive impairment are correlated with the severity of hearing loss resulting in a more rapid decline overtime (Acar et al., 2011; Amieva et al., 2015; Deal et al., 2015; F. R. Lin et al., 2013; F. R. Lin, Ferrucci, et al., 2011; F. R. Lin, Thorpe, et al., 2011; Slade et al., 2020; Wayne & Johnsrude, 2015).

The use of a CI for those with severe-profound hearing loss, has shown improvements in cognitive ability, when comparing pre-versus post-operative outcomes, specifically in terms of learning, memory, language, executive functioning and attention (Ambert-Dahan et al., 2017; Castiglione et al., 2016; Claes et al., 2018; Cosetti et al., 2016; Mertens et al., 2020; Mosnier et al., 2015, 2018; Sarant et al., 2019; Sonnet et al., 2017) and in quality of life measured across various different tests and domains such as emotional well-being, physical, emotional and social role functioning, mental health and vitality (Arnoldner et al., 2014; Chung et al., 2012; Cohen et al., 2004; Contrera et al., 2016; Damen et al., 2007; Djalilian et al., 2002; Labadie et al., 2000; Maillet et al., 1995; Ramos-Macías et al., 2016; Rostkowska et al., 2021; Sarant et al., 2019; Sonnet et al., 2017; Waltzman et al., 1993).

When investigating types of CI users, bilateral CI users show more benefits in speech perception, localization of sound and speech, and cognition compared to unilateral only and unilateral with a hearing aid (unilateral/HA) user (Blamey et al., 2015; Hua et al., 2017; Loizou et al., 2009; Noble et al., 2008; A. Schäfer et al., 2011; van Hoesel & Tyler, 2003). The use of bilateral CIs allows for more accurate sound localization and therefore, and in a social setting, this can improve SNR because of the ability to orient themselves towards the talker is better (van Hoesel, 2015) and a release from masking (Mok et al., 2007). The implantation of a second CI, therefore, may reduce cognitive burdens of listening as shown by lower listening effort reports by bilateral CI users compared to unilateral/HA and unilateral only (Noble et al., 2008; Schnabl et al., 2015). It is also shown through differences in visual working memory performance between unilateral and bimodal users and scores were also correlated with speech perception in quiet only in those with bimodal listening (Hua et al., 2017).

However, compared to normal hearing (NH) individuals, in one study, CI users, in general, tend to exhibit a lower degree of cognitive ability, in terms of immediate memory, visuospatial skills, language, attention and delayed memory (Claes et al., 2018). In another study, CI users exhibited lower performance scores tests measuring in global cognition, verbal episodic memory, processing speed, attention, and cognitive flexibility (Huber et al., 2020). One study also showed lower reports of quality of life in CI users compared to NH controls more significantly in specific sections of the World Health Organization Quality of Life Group (WHOQOL) test (Fleck et al., 2000) pertaining to physical security, home environment, financial resources, health care, access to information, recreation and leisure, physical environment and transportation (de Angelo et al., 2016). These suggest that the use of a CI might not completely recover cognitive ability and quality of life outcomes to the level of NH individuals and that some individuals still present or develop mild cognitive impairment later in life despite the benefits of using a CI (Mosnier et al., 2018).

In this present study, we used online testing and surveys to measure working memory ability in the visual, auditory in quiet and auditory in noise conditions, speech perception in quiet and noise, quality of life and quality of hearing in CI users. NH controls were also recruited to perform the working memory and speech perception tests along with quality of hearing. The objective of this study is to investigate the effects of different hearing and listening conditions on cognition and working memory and see if those results, along with demographic variables, can predict clinical speech perception and subjective reports of quality of life and quality of hearing. Also, we would like to demonstrate that online testing can be used as a tool to assess hearing, cognition, quality of life and quality of hearing in CI users. For this study, we tested the following hypotheses: (1) CI users’ online test performance and survey outcomes will be significantly lower than NH controls, (2) bilateral CI users will perform better on the online tests and report higher outcomes on the surveys compared to unilateral/HA users and unilateral only users, (3) and unilateral/HA users would perform better compared to unilateral only, (4) performance on the visual working memory test will be significantly greater than auditory in quiet and in noise, (5) performance on auditory working memory and speech perception in quiet will be significantly greater than in noise, (6) CI users with greater online test scores will also show greater speech perception and better reports of quality of life and quality of hearing, and (7) performance on online tests will predict speech perception, and sections of quality of life and quality of hearing outcomes.

## 2. Methods

### 2.1. Participants

Demographic information of all participants is shown in Table 1. Thirty-one CI users were recruited from the patient population in the Department of Otolaryngology at Sunnybrook Health Sciences Centre. Ages ranged between 20 and 82 years (M = 53.5, SD = 19.1) and included seventeen males and fourteen females with no underlying neurological conditions. CI users consisted of ten bilateral users, twelve unilateral users, and nine unilateral users with a contralateral HA. Demographics also include speech perception in quiet and in noise measured by AzBio tests (Spahr & Dorman, 2005) conducted as apart of their standard clinical testing. Also included are hearing condition (bilateral, unilateral/HA and unilateral only), duration of implantation (DOI), age of implantation (AOI; range = 2-80, M = 44.3, SD = 24.5), and duration of deafness (DOD), which was obtained by subtracting the date of implantation from their subjective reports on when their hearing loss began. Age and AOI was not included in Table 1 for the privacy of participants. These variables for each participant were used for correlational analyses along with outcomes from online tests and surveys they completed. Thirty-one age-matched controls were also recruited with ages ranging from 20-85 (M = 53, SD = 17.2) and included fourteen males and seventeen females with no underlying neurological conditions. They were recruited through local databases and online social media groups in the Toronto, Canada area.

**Table 1:**
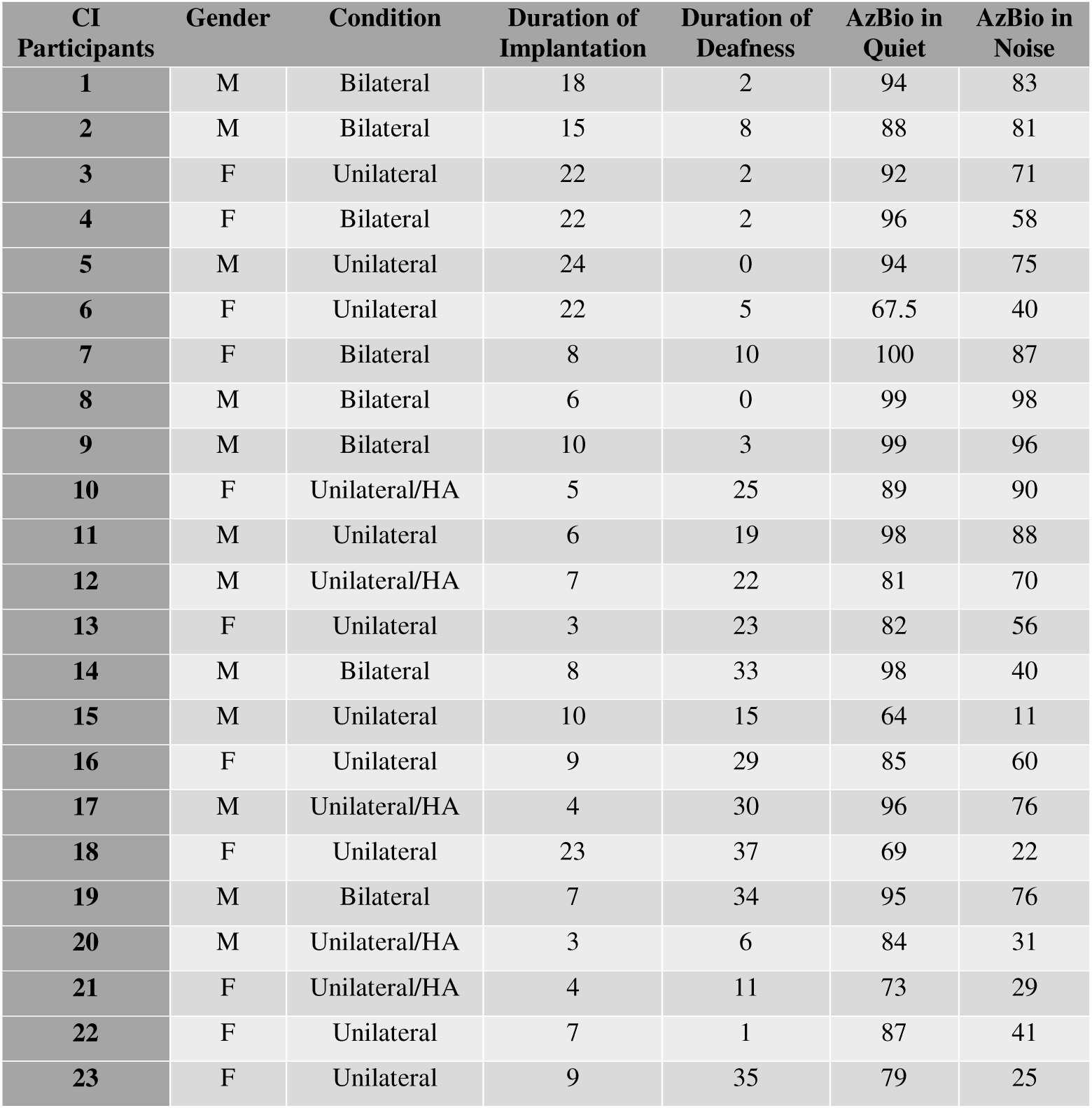
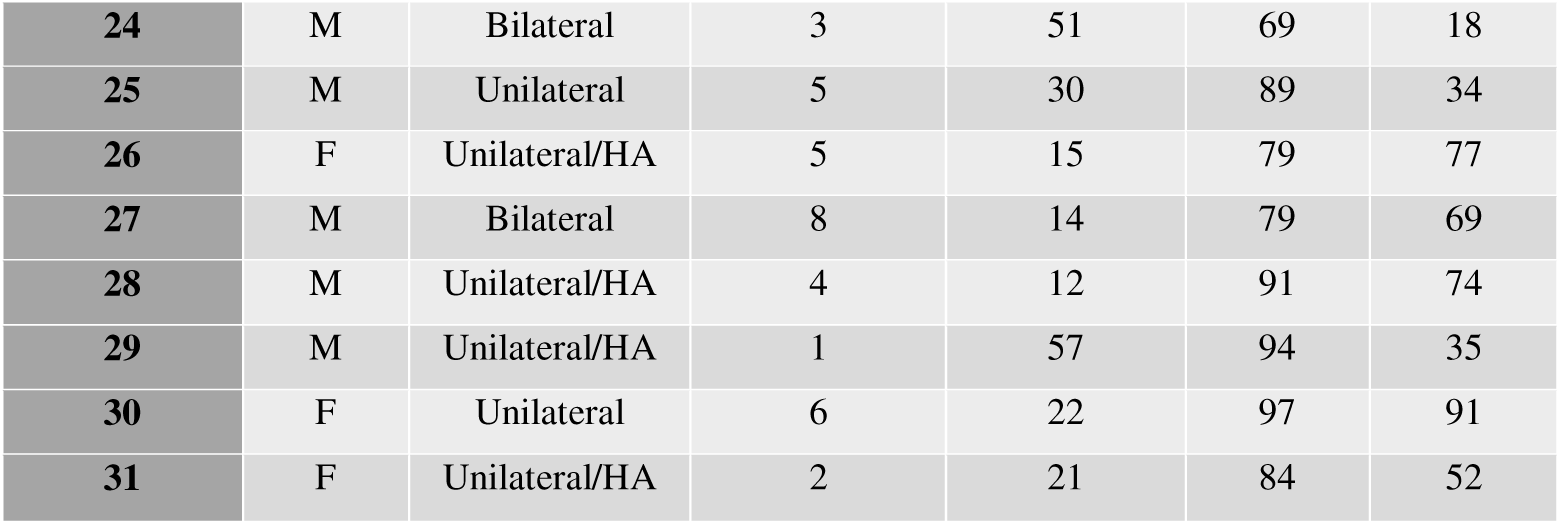
Demographic of Participants. Table at the top are CI participants participants. The individuals are numbered 1 to 31 from youngest to oldest and their corresponding condition (bilateral, unilateral, and/or if a HA, is used), duration of CI use in years, duration of deafness before implantation and their outcomes of their speech perception tests (AzBio) are recorded, respectively, in the columns from left to right.

All participants provided written and informed consent for the study procedures, which were conducted in accordance with the Research Ethics Board (REB) at Sunnybrook Health Sciences Centre. The approved protocol was in agreement with the Declaration of Helsinki. Participants were monetarily compensated for their participation and were provided full reimbursement for parking fees at the hospital campus.

### 2.2. Online Tests and Surveys

All online tests and surveys were created using JavaScript and uploaded onto an online server. Participants were tasked with three working memory sentence span tasks (Conway et al., 2005; Daneman & Carpenter, 1980): reading span, listening span in quiet and listening span in noise, each following a similar paradigm but in different listening conditions and completed matrix sentence tests to measure speech perception in quiet and noise (Wagener et al., 1999). Before starting the auditory tests, participants are given the opportunity to adjust their volumes to a comfortable level. Two surveys were also completed measuring quality of different aspects of life and hearing. Since the tasks are available online, participants performed all on their personal mobile devices or computers.

### 2.3. Working Memory Tasks

In all three sentence span tests, participants were presented with a sentence and asked to make a judgement on if the sentence made sense. An example of the trial structure is shown in Figure 1A for reading span and Figure 1B for both listening span tests. The sentence for reading span is shown until decision is made and during the listening spans, the sentence is played once with the listening span in noise being played with a multi-talker babble in the background at a signal-to-noise ratio (SNR) of +10. After pressing “Yes” or “No”, a single word was visually presented for 950ms and removed from screen for reading span. For the listening span tasks, the words are auditorily presented for around 950ms, depending on the length of the word, with multi-talker babble in the background in the noisy condition. Participants are asked to memorize these words in order and recall it later. Each set contains varying amounts of sentence-word pairings from two to six and once presented, participants are asked to recall all the words; for all five set sizes, each set size is presented a total of three times and therefore, participants undergo fifteen trials in total with two trials beforehand for practice. Performance on each task is measured by the amount words correctly recalled across all set sizes.

**Figure 1:**
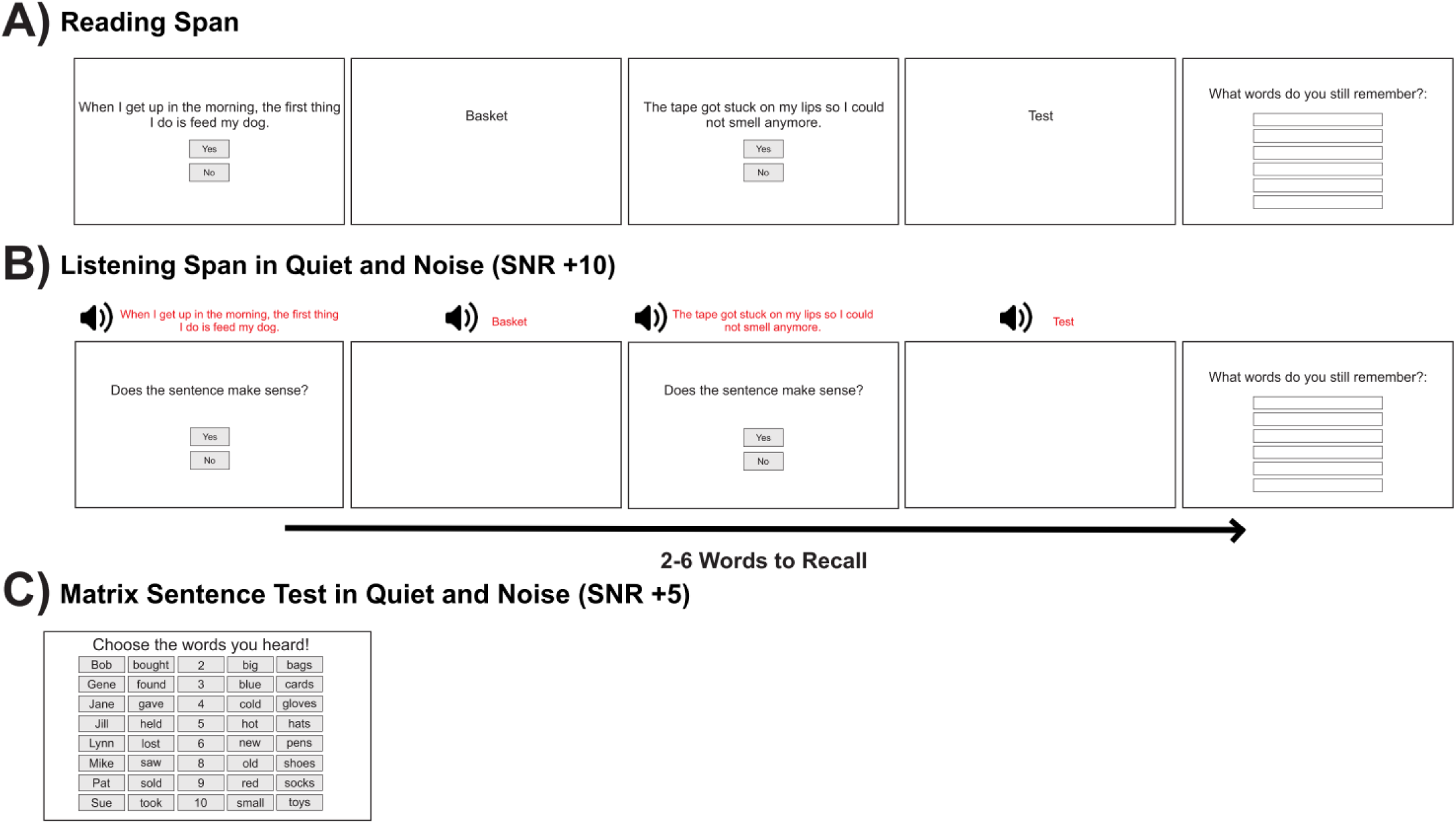
Schematic diagram of online tests. A) Reading span working memory test. A sentence is visually presented with a “Yes” and “No” button. Participant must identify if the sentence makes sense. Then a word is presented that must be remembered. This occurs in 2-6 sets. B) Listening span working memory tests. A similar paradigm to the reading span test however, the sentences and words are presented auditorily as indicated by the red text. It consists of two conditions with a quiet background and with a noisy background (SNR +10). C) Matrix sentence test. A simple 5-word sentence is presented auditorily and participants must listen and respond with the sentence that was heard. It consists of two conditions with a noisy background (SNR +5) and with a quieter background (SNR +20).

### 2.4. Matrix Sentence Tests

Participants completed two versions of the matrix sentence test with different levels of white background noise: SNR of +20 for the quieter condition and SNR of +5 for the noisy condition. They completed 10 sentences for each condition with optional practice trials before starting the test. Each sentence consisted of five, single syllabled words following the same format: person’s name, action in past tense, a number from two to nine excluding seven, an adjective, and an object (Figure 1C). Performance on both tasks were measured by number of words correctly identified across all sentences presented in that condition.

### 2.5. CIQOL-35 Survey

The CIQOL-35 profile instrument was tailored specifically for cochlear implant users (McRackan et al., 2017, 2018; McRackan, Hand, Velozo, & Dubno, 2019; McRackan, Hand, Velozo, Dubno, et al., 2019). The survey is comprised of 35 questions as a measure to assess the impact of their hearing ability on their functional ability in 6 different domains: communication (receptive and expressive communication ability in different situations), emotion (impact of hearing ability on emotional well-being), entertainment (enjoyment and clarity of TV, radio, and music), environmental (ability to distinguish and localize environmental sound), listening effort (degree of effort and resulting fatigue associated with listening), and social (ability to interact in groups and to attend and enjoy social functions).

The items are scored using the same five answers (never, rarely, sometimes, often and always) with each answer corresponding to a number from 1 to 5 or in some cases, the reverse. The scores are then summed for each domain and then converted to the interval-scale score as derived from item-response theory (McRackan, Hand, Velozo, & Dubno, 2019; McRackan, Hand, Velozo, Dubno, et al., 2019). The higher the score (from 0 to 100), the better their subjective functional ability is in the different sections measured.

### 2.6. SSQ Survey

This survey assesses three domains of hearing, speech hearing, spatial hearing and other qualities of hearing which includes the ability of individuals to segregate and recognize sounds, the clarity of speech, and listening effort (Gatehouse & Noble, 2004). The number of questions vary for each section with fourteen dedicated to speech hearing, seventeen for spatial hearing and eighteen for other qualities of hearing. Questions for each section are answered by a rating between zero and ten associated with the answers “Not at all” and “Perfectly”, respectively. The ratings are then averaged for each section. The higher the scores (from 0 to 10), the better their ability or hearing is, subjectively.

### 2.7. Statistics

In this paper, these outcomes from online tests and surveys as well as demographic variables mentioned above were all correlated against each other. For the hearing condition, each categorical variable was assigned a number; unilateral only CI users were “1”, unilateral with a HA was “2”, and bilateral CI users were “3”. Correlation analysis was performed using R (R Core Team, 2020) using the *psych* package to create a triangular correlational matrix in which it presented significant Spearman correlations all corrected for false discovery rate (FDR). In order to identify the most predictive online outcome measures and demographic variables to each different section of both surveys, partial least squares (PLS) regression was used (*caret* package). PLS was chosen due to the high amount of collinearity among variables in this study and its ability to identify predictors that contribute most to the model to maximally explain the outcome variable which can graphed as variable importance in the projection (VIP) plots. For each regression test, 75% of predictor and outcome variables were trained using leave one out cross validation (LOOCV). The model was then tested on the remaining 25% resulting in a root mean squared error (RMSE) and R-squared (R^2^) value indicating the accuracy of the model to predict the outcome. While RMSE is a metric showing how far the predicted values are compared to the original, R^2^ indicates the proportion of variance in the outcome variable that can be explained by the predictor variables. The variables contributing more than 50% to a model were chosen and inputted into a multivariable linear regression model.

T-tests were also performed on the online tests (sentence span and matrix tests) comparing performance in different conditions. T-tests were also performed on the online tests and SSQ survey outcomes between different groups. All t-tests and Pearson correlations were two-tailed, and the alpha criterion for Type I error was set at 0.05 with effect sizes for t-tests expressed by Cohen’s *d*.

## 3. Results

### 3.1. Behavioural Results

#### 3.1.1. Online Test Performance and Survey Outcomes

CI and NH performance for recall ability in the sentence span tests and matrix sentence tests are shown in Figure 2. The averages and standard deviations for each test for the CI group are 62.4% (23.2), 31.3% (19.1) and 19.6% (18.9) for recall performance in reading, listening in quiet and listening in noise, respectively and for the NH group, 57.3% (24.3), 58.9% (24.6) and 55.2% (21.4). For the matrix tests in quiet and in noise, the CI averages and standard deviations are 89% (9.9) and 79.4% (14.8) and for NH, 96.9% (5.7) and 94.3% (7.7). For the SSQ sections, the CI averages and standard deviations for speech, spatial and quality of hearing, respectively, are 5.66 (2.3), 4.66 (2.6) and 6.88 (2.3) and for NH, the scores are 7.47 (1.4), 7.46 (1.9) and 8.32 (1.4). Between group comparisons for each test showed no significant difference in reading span performance however, the CI group performed significantly worse on the listening span in quiet and noise (t(30) = -4.92, p < 0.0001, *d* = 1.25; t(30) = -6.94, p < 0.0001, *d* = 1.76) and in the matrix tests in quiet and noise (t(30) = -3.83, p = 0.0003, *d* = 0.95; t(30) = -4.97, p < 0.0001, *d* = 1.26). Subjective reports of speech, spatial and quality of hearing were significantly poorer in CI users as well compared to NH (t(30) = -3.77, p = 0.0004, *d* = 0.95; t(30) = -4.83, p < 0.0001, *d* = 1.23; t(30) = -2.95, p = 0.0045, *d* = 0.76, respectively).

**Figure 2:**
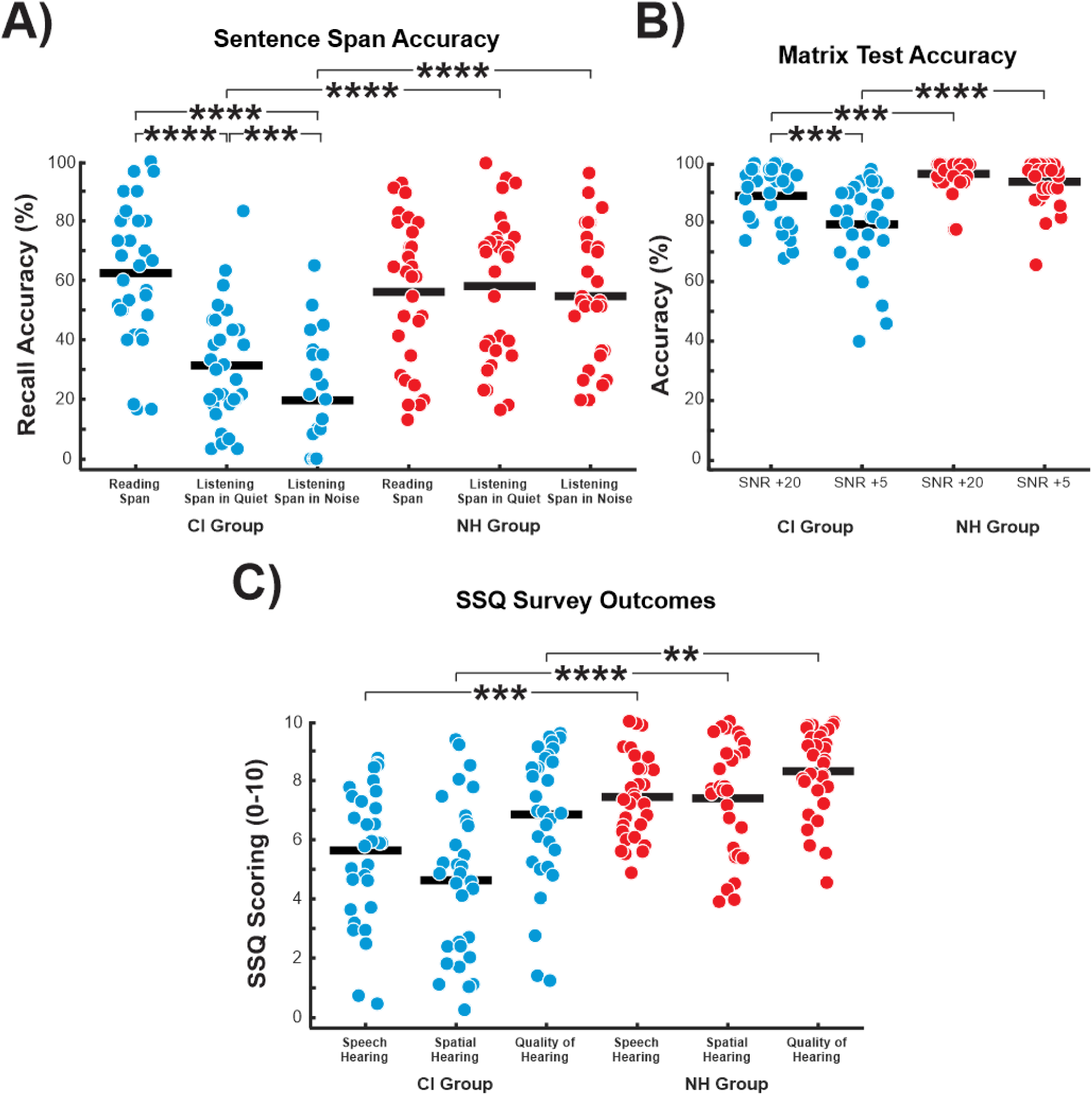
Performance on sentence span and matrix tests. A) Accuracy on recalling the sets of words in sentence span tests. CI users performed the best on the reading span, and this was significantly greater than listening span in quiet and listening span in noise. Scores on listening span in quiet were significantly greater than that in noise. B) Accuracy on recalling the simple sentences in matrix sentence tests. CI users performed better on the quieter condition compared to the noisy. **** = p < 0.0001, *** = p < 0.001, ** = p < 0.01

Within group comparisons showed that CI users performed better on the reading span test compared to both listening span in quiet and in noise in recall ability (t(30) = 7.95, p < 0.0001, *d* = 1.46; t(30) = 9.81, p < 0.0001, *d* = 2.02) and they performed better on listening span in quiet compared to in noise (t(30) = 4.06, p = 0.0003, *d* = 0.62). This was also seen in the matrix sentence tests where performance was higher in the quiet condition than in the noisy (t(30) = 5.13, p < 0.0001, *d* = 0.92). In the NH group, no significant differences were found between performances of each working memory and speech perception online tests however, descriptively, we find that NH participants performed similarly on the listening span in quiet compared to reading span and performed the worst on listening span in noise.

#### 3.1.2. Hearing condition and online tests and surveys

The relationship between hearing condition (bilateral, unilateral/HA or unilateral only) and online tests were investigated. This was tested because past studies have found the relationship to be significant where those with bilateral implantation report benefits in speech perception (Laszig et al., 2004; Müller et al., 2002; Neuman et al., 2007; Noble et al., 2009; Verschuur et al., 2005). This study corroborated those findings and are shown in Figure 3; bilateral CI users performed better than the unilateral only group on all three working memory tasks (reading span: t(20) = 2.37, p = 0.028, *d* = 1.00; listening span in quiet: t(20) = 3.31, p = 0.0035, *d* = 1.40; listening span in noise: t(20) = 2.10, p = 0.048, *d* = 0.88) and on the two speech perception tasks (matrix test in quiet: t(20) = 2.61, p = 0.017, *d* = 1.23; matrix test in noise: t(20) = 2.78, p = 0.012, *d* = 1.14). Compared to the unilateral with a HA group, bilateral CI users only outperformed them on the listening span in quiet and matrix test in quiet (t(17) = 2.57, p = 0.02, *d* = 1.20; t(17) = 2.45, p = 0.025, *d* = 1.11, respectively); on the other tests, the average indicated a trend for a similar result however, it was not significant. Comparison between unilateral with a HA and unilateral only group resulted in no significant differences.

**Figure 3:**
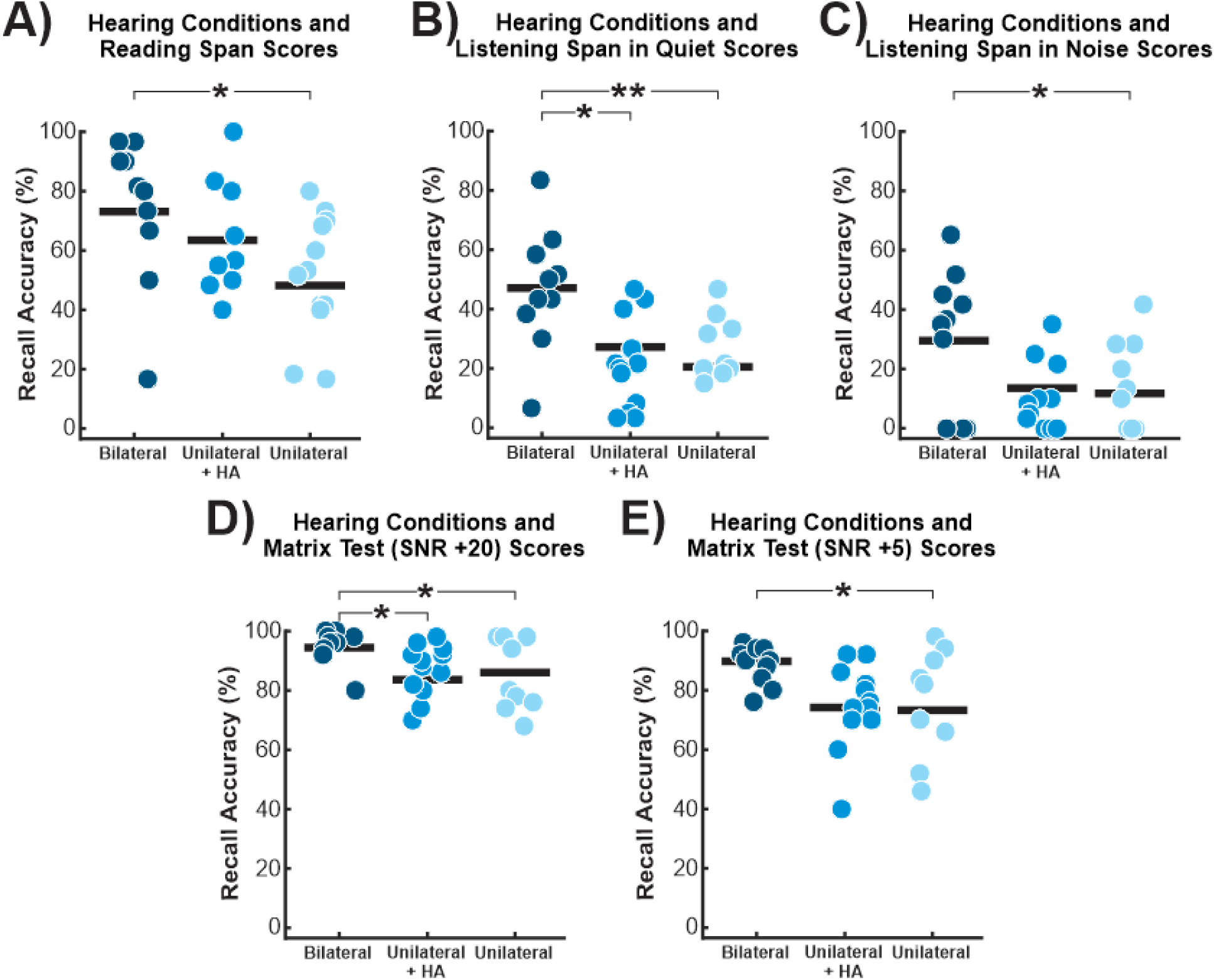
Performance on online tests between each hearing conditions. Comparison of performance between hearing conditions in CI users on A) reading span, B) listening span in quiet, C) listening span in noise, D) matrix test in quiet, and E) matrix test in noise. ** = p < 0.01, * = p < 0.05.

Multiple linear regression on each of the online tests with the demographic variables were assessed. Included in the model was age, duration of deafness, duration of implantation, and hearing condition; age of implantation was removed from the models due to its high variance inflation factor. For listening span in quiet, the model was significant and predicted 33% of variability in scores (p = 0.006), also for listening span in noise it predicted 20% of its variance (p = 0.043). For matrix test in quiet the model predicted 21% of variance (p = 0.037) and for matrix test in noise it predicted 28% of variance (p = 0.014). Hearing condition was a significant predictor for listening span in quiet (β = -10.8, SE = 3.72, F(1,27) = 8.43, p = 0.007) and matrix test in quiet (β = -4.62, SE = 2.09, F(1,27) = 4.90, p = 0.036). These suggest that there’s a decrease of 10.8% in listening span in quiet scores and a decrease of 4.62% in matrix test in quiet scores when it goes from bilateral to unilateral/HA users or from unilateral/HA to unilateral only. Age was a significant predictor for matrix test in noise (β = -0.44, SE = 0.21, F(1,27) = 10.7, p = 0.003) suggesting that there’s a decrease of 0.44% in matrix test in noise scores when there’s an increase of 1 year in age. The model for reading span was not significant.

Figure 4 shows the comparison of hearing conditions in each section of the CIQOL and SSQ surveys and AzBio in quiet and noise scores. Significant differences were found in the CIQOL entertainment section where bilateral CI users reported better enjoyment and clarity of media compared to both unilateral/HA users (t(17) = 2.88, p = 0.01, *d* = 0.90) and unilateral only (t(17) = 2.11, p = 0.047, *d* = 0.62). Bilateral CI users also reported lower levels of listening effort compared to unilateral/HA users (t(17) = 2.36, p = 0.03, *d* = 1.07) however, no significant difference was found with unilateral only. For the SSQ survey, bilateral CI users significantly reported better speech, spatial and quality of hearing compared to unilateral/HA users (t(17) = 2.70, p = 0.02, *d* = 1.22; t(17) = 2.84, p = 0.01, *d* = 1.28; t(17) = 3.06, p = 0.007, *d* = 1.38) and compared to unilateral only, only reported better scores on speech and spatial hearing (t(17) = 2.72, p = 0.01, *d* = 1.18; t(17) = 6.09, p < 0.0001, *d* = 2.61).

**Figure 4:**
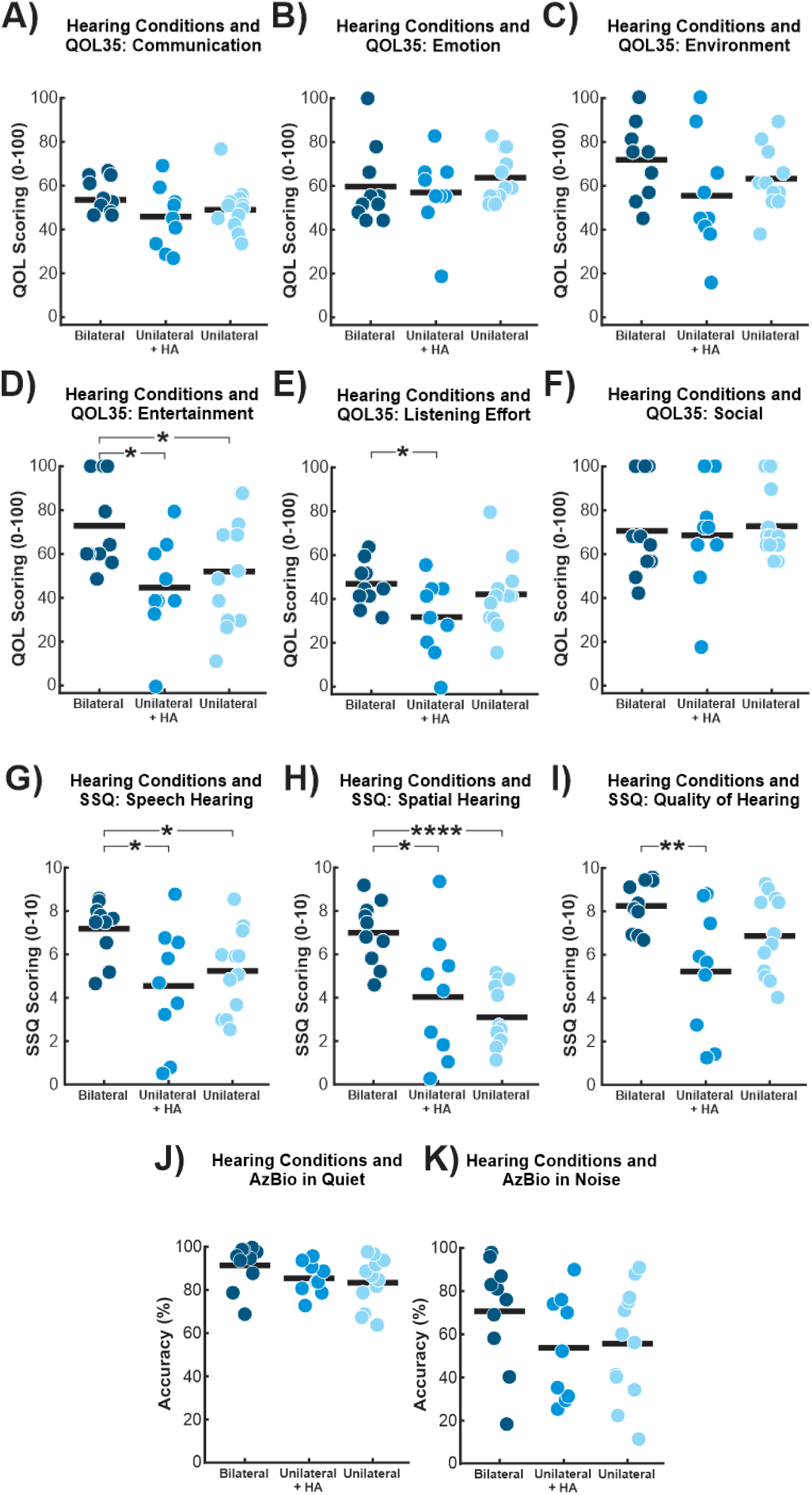
Comparison of hearing conditions and CIQOL and SSQ survey outcomes. Quality of life and hearing scores for each individual after completing all six sections of CIQOL: A) communication, B) emotion, C) environment, D) entertainment, E) listening effort, and F) social and all three sections of SSQ: G) speech, H) spatial, and I) quality of hearing, J) AzBio in quiet and K) AzBio in noise. **** = p < 0.0001, ** = p < 0.01, * = p < 0.05

### 3.2. Correlation Results

#### 3.2.1. CI users

Performance on the online tests, survey outcome scores for each section, clinical speech perception scores and demographic variables (age, age of implantation, duration of deafness before implantation, duration of implantation) were all used for correlational analysis. Figure 5 shows the correlational triangular matrix result where only significant correlations that survived multiple comparisons correction by FDR, are present. Overall, the online test performance scores show significant positive correlations with certain sections of CIQOL and SSQ as well as AzBio clinical scores. Certain demographic variables also show significant correlations with sections of CIQOL and the online tests. For hearing condition, a number was assigned to each condition where unilateral only, unilateral/HA and bilateral users were numbers “1”, “2”, and “3”, respectively.

**Figure 5:**
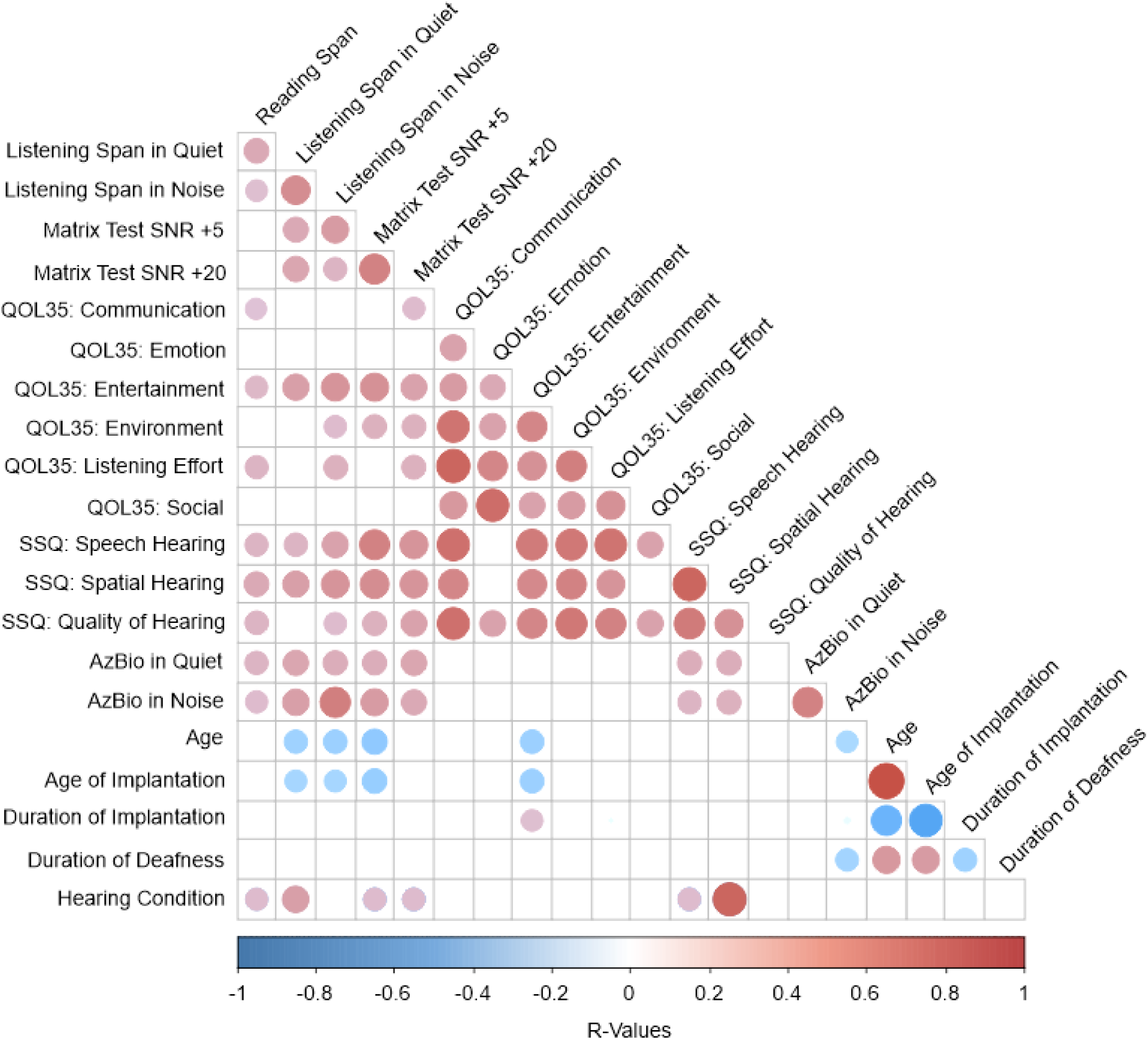
Correlation matrix CI outcomes. Each circle represents a significant correlation after correcting for multiple comparisons using FDR. Red circles indicate a positive correlation while blue indicates a negative correlation. The bigger and darker the circle, the greater the correlation.

The CIQOL sections positively correlated with the online tests are communication with reading span and matrix test in quiet, entertainment for all five online tests, environment for listening span in noise and matrix test in quiet and in noise, and listening effort for reading span, listening span in noise and matrix test in quiet. The speech and spatial hearing sections in the SSQ survey are positively correlated with all five tests and the other qualities of hearing section was positively correlated with reading span, listening span in quiet and in noise and matrix test in quiet and in noise. All five online tests were also positively correlated with AzBio in quiet and AzBio in noise. In terms of demographics, age and age of implantation were negatively correlated with the entertainment section from CIQOL and duration of implantation was positively correlated. Age and age of implantation were also negatively correlated with listening span in quiet, in noise and with matrix test in noise. Age and duration of deafness were found to be negatively correlated with AzBio in noise. Hearing condition was positively correlated to reading span, listening span in quiet, and matrix tests in quiet and noise along with positively correlated to the speech and spatial sections of SSQ suggesting that bilateral CI users performed better and reported higher scores on the tests and surveys.

#### 3.2.2. NH controls

Performance on the online tests, SSQ survey outcome scores for each section, and age were all used for correlational analysis. Figure 6 shows the correlational triangular matrix result for the NH groups; shown are significant correlations that survived multiple comparisons correction by FDR. Overall, the online working memory performance scores, all three tests, show significant positive correlations with only the spatial section of SSQ while age was significantly and negatively correlated with the speech and spatial section of SSQ.

**Figure 6:**
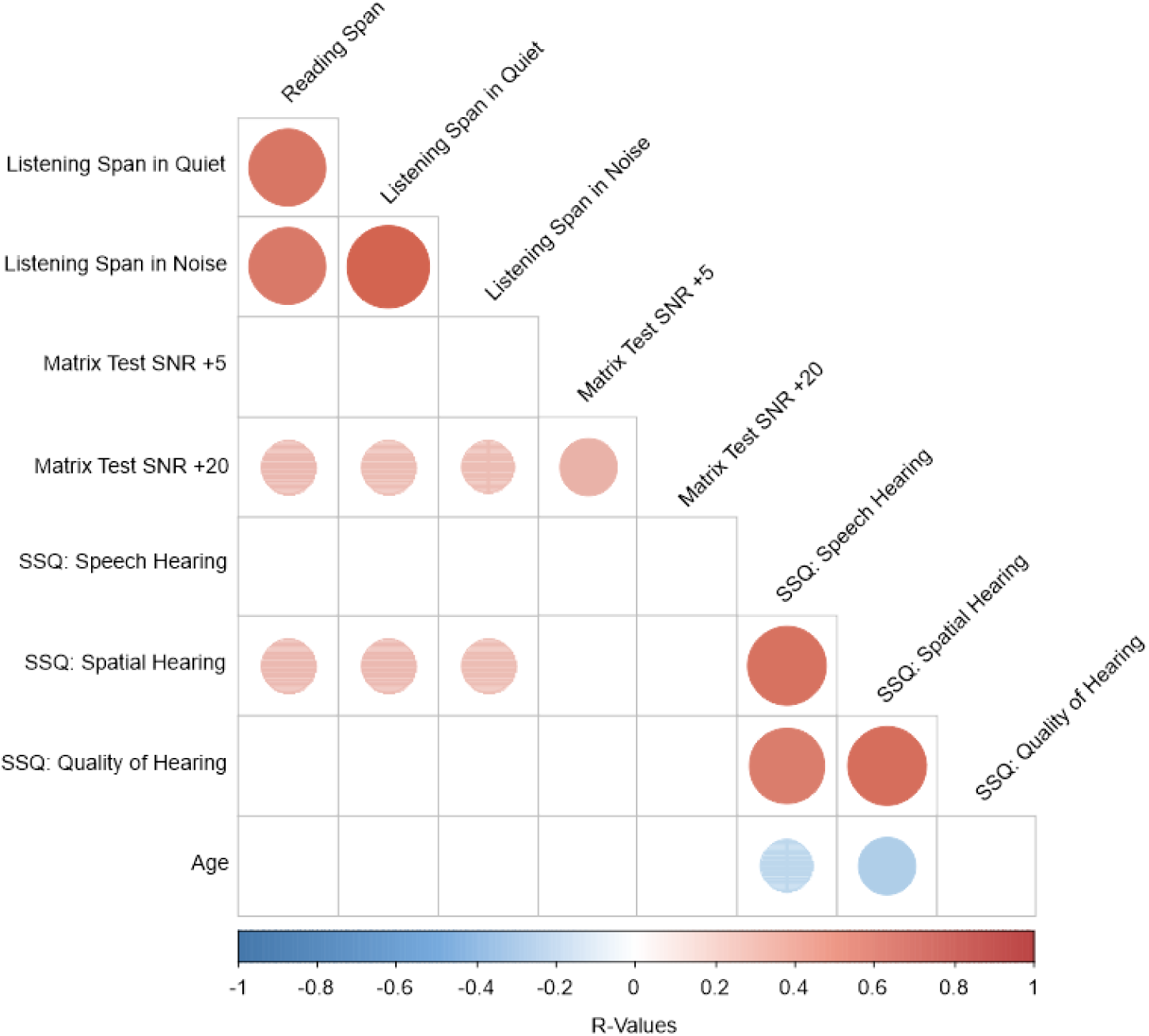
Correlation matrix NH outcome. Each circle represents a significant correlation after correcting for multiple comparisons using FDR. Red circles indicate a positive correlation while blue indicates a negative correlation. The bigger and darker the circle, the greater the correlation.

### 3.3. Regression Results

#### 3.3.1. CI Users

##### 3.3.1.1. Partial Least Squares (PLS) Regression

Figure 7 shows results from the PLS regression analysis. The performance on online tests and demographic variables were used to predict survey outcomes in each section along with AzBio in quiet and in noise performance where the bar plots indicate a hierarchy of variables from most contributing to least for each survey section and clinical speech perception outcome. The table in Figure 7 shows R^2^ and RMSE results from each analysis in which the model contained all online tests and demographic variables.

**Figure 7:**
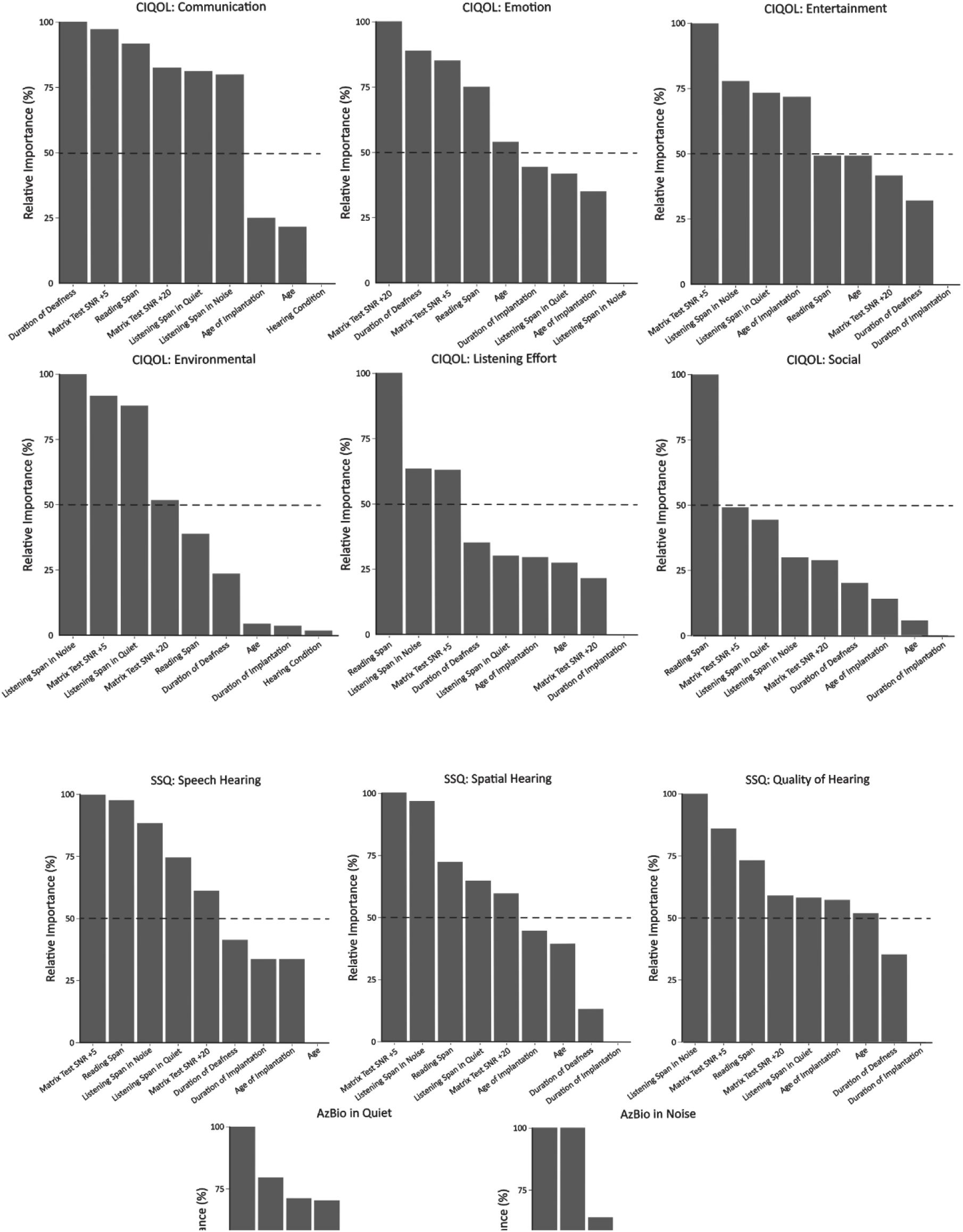
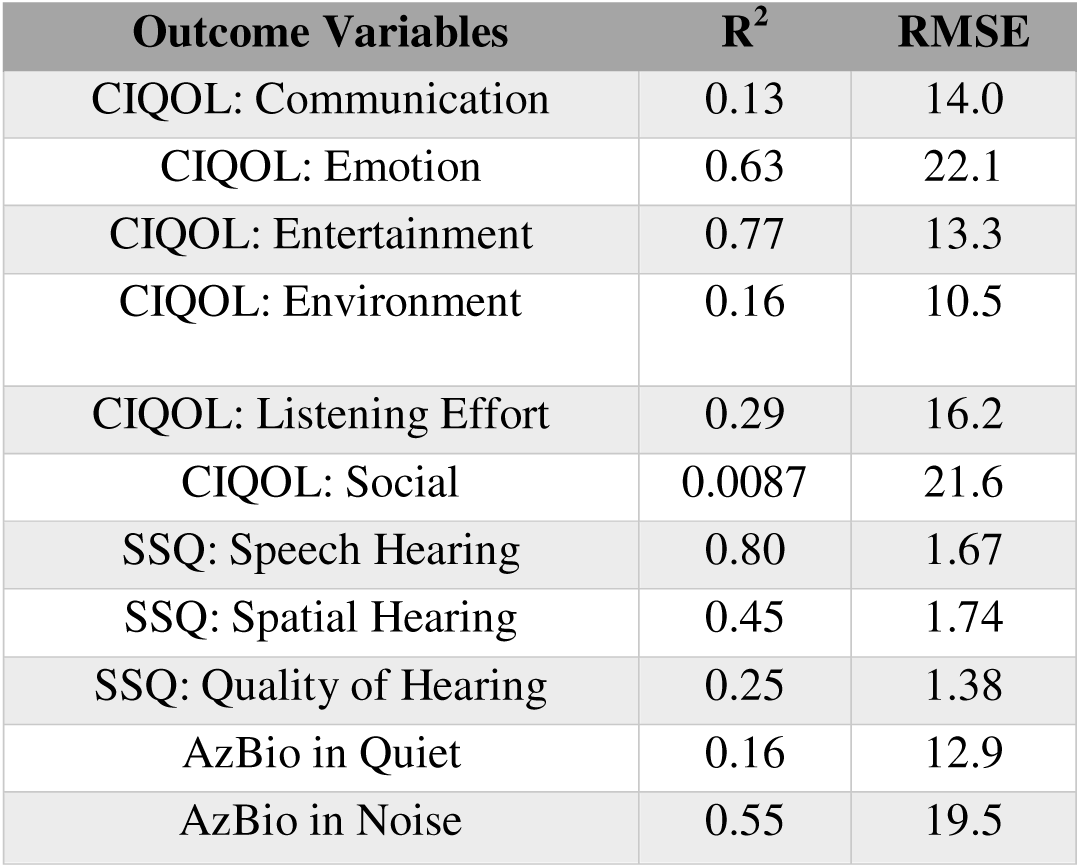
Relative Importance Plots in CI users. Feature selection is performed on each outcome variable to predict them using online test performance and demographic variables. The relative importance bar plots for each outcome variable are plotted where variables contributing over 50% are indicated by the horizontal dotted line. The table at the bottom indicates the outcome variables for prediction on the left column and on the right columns are the R^2^ and RMSE values, respectively, for each model.

##### 3.3.1.2. Multiple Linear Regression Results

Using the PLS results above, for each outcome variable, the variables contributing more than 50% were obtained and multiple linear regression was performed in order to explain the variability of the survey and speech perception outcomes; variables that exhibited a VIF value of 3 or more were excluded. According to the linear regressions performed, the online tests and demographics were able to significantly predict the entertainment section of CIQOL, speech, spatial and quality of hearing from the SSQ survey and AzBio in noise.

One of the significant models was the prediction of clinical AzBio in noise scores, which measured speech perception in noise, where the model included listening span in noise, duration of deafness, and duration of implantation. The results showed that the overall model significantly accounted for ∼59% of variance in AzBio in noise (p < 0.0001) and significant predictors included listening span in noise (β = 0.92, SE = 0.16, F(1,27) = 38.57, p < 0.0001) and duration of deafness (β = -0.61, SE = 0.22, F(1,27) = 5.02, p = 0.01). These suggest that, for listening span in noise, there’s an increase of 0.92% in AzBio in noise scores when there’s an increase of 1% in listening in noise recall performance and, for duration of deafness, there’s a decrease of 0.61% in AzBio in noise scores when there’s an increase of 1 year in duration of deafness. When assessed as isolated Spearman correlations, the correlation between AzBio in noise and listening span in noise was 0.73 (p < 0.0001; Figure 8A) while the correlation between AzBio in noise and duration of deafness was -0.41 (p = 0.04; Figure 8B).

**Figure 8:**
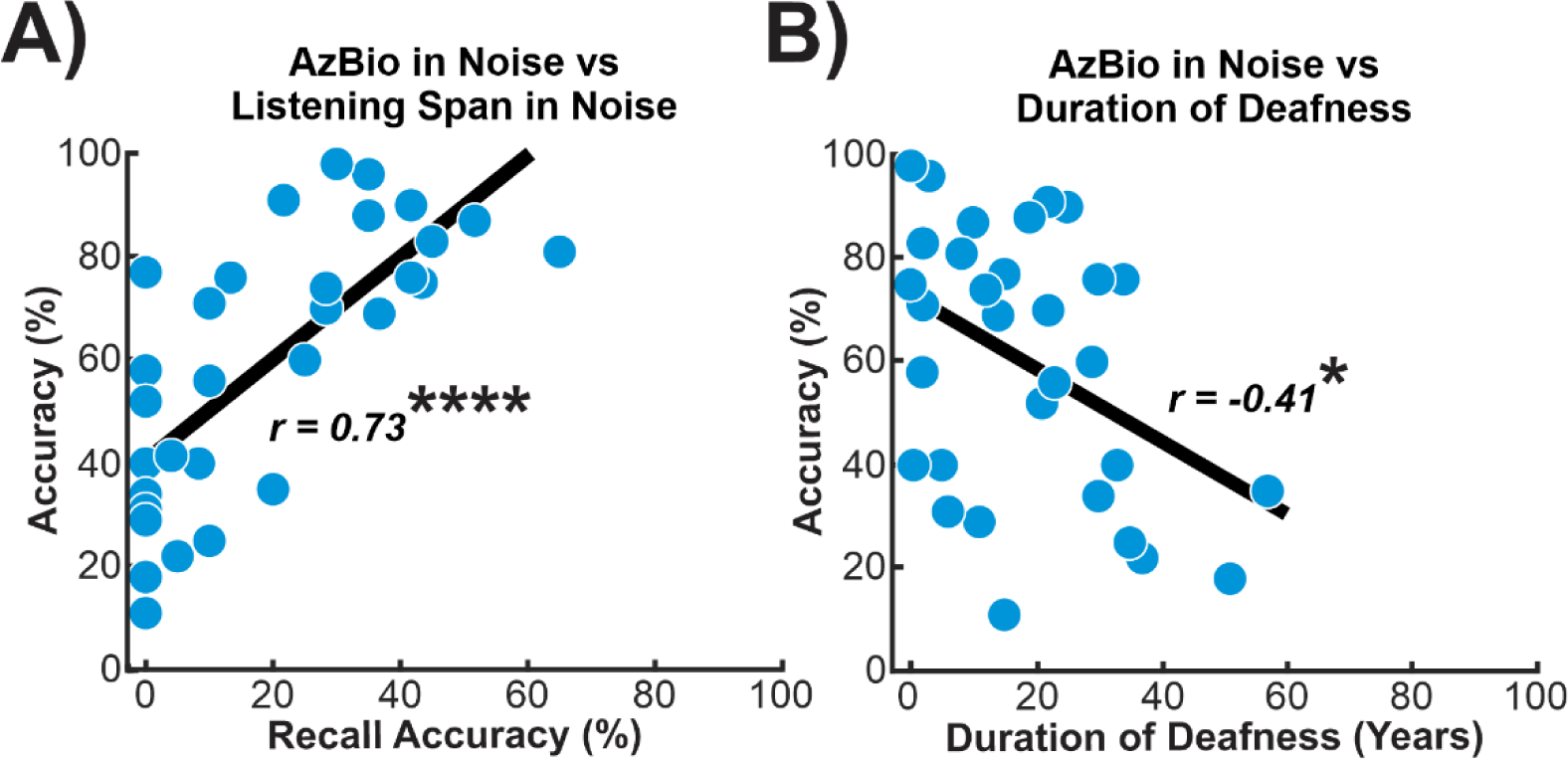
AzBio in noise correlation results. A) Correlation between AzBio in noise scores and recall performance on listening span in noise. B) Correlation between AzBio in noise scores and duration of deafness. **** = p < 0.0001, * = p < 0.05.

Other significant results included models predicting the speech, spatial and quality of hearing sections in SSQ and the entertainment section of the CIQOL. For all four results, the model included all three sentence span tests and the two matrix sentence tests. The model for the speech hearing section significantly accounted for ∼48% of variance in subjective speech hearing scores (p < 0.0005), for spatial hearing, the variables significantly accounted for ∼50% of variance in subjective spatial hearing (p < 0.0005) and for quality of hearing, the variables significantly accounted for ∼21% of variance in subjective ease of listening, clarity and differentiation of different speaker. The model for the entertainment section significantly accounted for ∼42% of variance in subjective enjoyment and clarity of media (p = 0.002). However, no significant predictors were found for all models.

#### 3.3.2. NH Controls

##### 3.3.2.1. Partial Least Squares (PLS) Regression

NH group performance on online tests and age were used to predict SSQ survey outcomes in each section. Figure 9 shows the bar plots indicate a hierarchy of variables from most contributing to least for each survey section and the table in shows R^2^ and RMSE results from each analysis in which the model contained all online tests and demographic variables.

**Figure 9:**
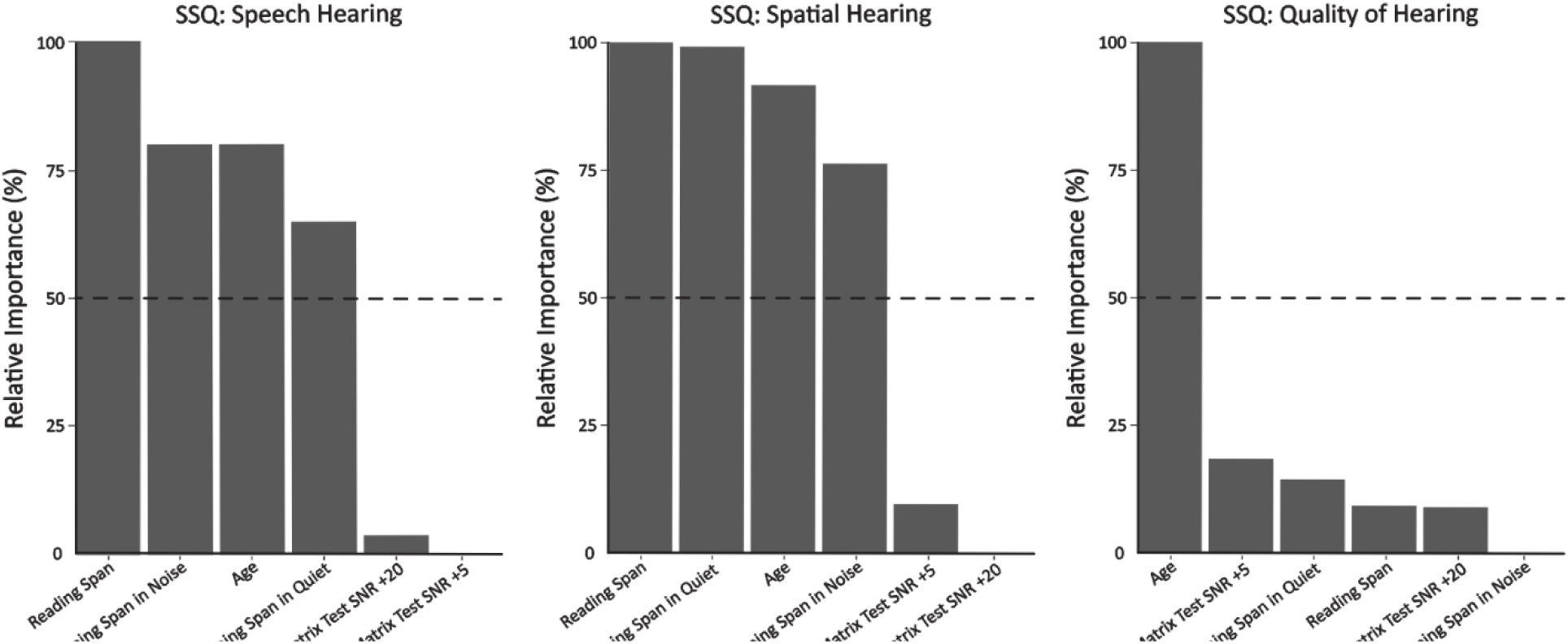
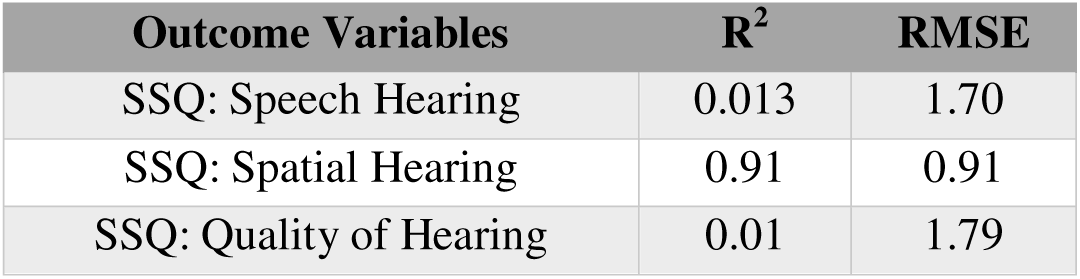
Relative Importance Plots in NH controls. Feature selection is performed on each outcome variable to predict them using online test performance and age. The relative importance bar plots for each outcome variable are plotted where variables contributing over 50% are indicated by the horizontal dotted line. The table at the bottom indicates the outcome variables for prediction on the left column and on the right columns are the R^2^ and RMSE values, respectively, for each model.

##### 3.3.2.2. Multiple Linear Regression Results

For each outcome variable, the variables contributing more than 50% were obtained from the PLS results and multiple linear regression was performed similar to the analysis done above with CI users. According to the linear regressions performed, the online working memory tests and NH ages (listening span in quiet was removed due to a large VIF) were only able to significantly predict the spatial section of SSQ accounting for ∼24% of variance in spatial hearing outcomes (p = 0.016) however, no significant predictors were found.

## 4. Discussion

### 4.1. Summary

The objective of this study was to investigate the effects of individual’s cognition and working memory on speech perception and subjective reports of quality of life and quality of hearing. The summary of results for this study are as follows: Hypothesis (1) was supported where performance on all online tests and listening conditions along with SSQ survey outcomes were significantly lower in CI users compared to NH controls. Hypothesis (2) was supported where bilateral users scored higher compared to the unilateral only group on online tests, reported greater SSQ survey outcomes on speech and spatial hearing sections and reported greater CIQOL survey outcomes only on the entertainment. However, compared to unilateral/HA, they only performed higher on the listening span and matrix test in quiet tasks and for survey outcomes, they reported higher scores on all sections of SSQ and the listening effort section on CIQOL. Hypothesis (3), however, was not supported where in unilateral/HA users performed and reported similar online and survey outcomes. Hypothesis (4) was supported where in CI users, performance on the visual working memory test was significantly greater than auditory in quiet and in noise. Hypothesis (5) was supported where in CI users, performance on auditory working memory and speech perception in quiet was significantly greater than in noise. Hypothesis (6) was supported where in CI users with greater online test scores, showed greater speech perception and better reports of quality of life and quality of hearing in specific sections. This is seen mainly, in CIQOL, communication, entertainment, environment, and listening effort sections and in SSQ, all three sections. Also, we found negative correlations with age and age of implantation with the entertainment section of CIQOL and the three online working memory tests along with a positive correlation between duration of implantation and the entertainment section of CIQOL and negative correlations between age and duration of deafness with speech perception in noise. Hypothesis (7) was supported where CI users’ performance on online tests predicted speech perception, quality of life and quality of hearing outcomes. AzBio in noise was independently predictable by listening span in noise and duration of deafness. Speech, spatial and quality of hearing sections of SSQ and the entertainment section of the CIQOL were predictable by all five online tests. Overall, these results suggest an impact of working memory and demographic variables to speech perception, quality of life and quality of hearing.

### 4.2. Working memory performance and listening conditions

#### 4.2.1. CI vs NH

In this study, when comparing CI users to NH controls, results show that, in the working memory tests, when verbal stimuli is presented in the visual modality, both groups perform similarly on recall however, when presented auditorily in both quiet and in noise, their performance is significantly worse. The similar performance in visual, verbal working memory is documented in other studies where CI users were shown to perform on par with NH controls (Lyxell et al., 2003; Moberly et al., 2016; Moberly, Harris, et al., 2017; Moberly, Houston, et al., 2017; Moberly, Pisoni, et al., 2017; O’Neill et al., 2019; Prince et al., 2021) suggesting that the phonological representations of words are intact and accessible in CI users. Results then show a decline in working memory recall performance once verbal stimuli are presented auditorily in quiet and only in CI users suggesting that, due to listening through a CI, working memory recall becomes affected (Moberly, Harris, et al., 2017; Moberly, Houston, et al., 2017); whether the decline is due to working memory ability or perception of the word presented is unclear in this study. Recall performance was impaired further when stimuli was presented auditorily with multi-talker babble in the background and this is shown through the working memory tests and matrix sentence tests. This is evident not only in CI users but also in NH controls, though, the decrease in average was larger in CI users suggesting perhaps a larger deficit in working memory ability when distractors are introduced.

A possible reason is that CI users allocate more attentional and working memory resources compared to NH during the encoding of visual, verbal stimuli (Lazard et al., 2010a; Prince et al., 2021) but is not reflected in behavioural performance then once auditory stimuli is introduced and they require more resources to disambiguate speech (Ala et al., 2020; Davis & Johnsrude, 2007; Miles et al., 2017; Rodd et al., 2005; Song et al., 2015), the limit to their working memory capacity is reached and performance may decline (Arlinger et al., 2009; F. R. Lin, Metter, et al., 2011; Pichora-Fuller et al., 2016; Pichora-Fuller & Singh, 2006; Rudner et al., 2011). Results are corroborated by other studies in which the addition of noise in the background elicits more cognitive resources to suppress the distracting stimuli (Cartocci et al., 2018; Dimitrijevic et al., 2017; Hjortkjær et al., 2020; McMahon et al., 2016; Petersen et al., 2015; Schneider et al., 2021; Wöstmann et al., 2015). This deficit from hearing loss or listening through a CI, and subsequent increased use of resources, can possibly affect quality of hearing as well, shown in this study (Hua et al., 2017; Kocak Erdem & Ciprut, 2019; Noble et al., 2008; Schnabl et al., 2015; Summerfield et al., 2006). When comparing SSQ survey outcomes between the two groups, we find that NH participants are more superior in all three sections where they report better speech perception, spatial hearing and quality of hearing which includes ease of listening along with clarity and identifiability of different speakers and sounds. This is addressed further below.

In addition to performance and survey outcome differences between groups, we also find that the two groups differ in which modality results in superior recall performance. For CI users, their visual working memory performance, compared to auditory, is the greatest and for NH controls, visual working memory and auditory working memory in quiet were comparable. In the case of CI users, due to their long term dependence on visual stimuli even after obtaining a CI, their visual working memory can be superior compared to auditory (Anderson et al., 2017; Bishop & Miller, 2009; Land et al., 2016; Lomber et al., 2010; Merabet & Pascual-Leone, 2010; Rouger et al., 2008; Song et al., 2015; Tye-Murray et al., 2007). However, considering that their visual working memory performance was equivalent to NH, it might be the case that access to their phonological storage through visual input, is still intact but once the stimuli is presented auditorily, CI users have difficulty accessing it and therefore, the use of increased working memory resources to attempt to disambiguate it, may result in detriments to performance.

#### 4.2.2. Bilateral vs Unilateral/HA vs Unilateral Only

We investigated the effects of hearing condition on working memory and speech perception and results show that bilateral users show superior performance compared to unilateral only in all five tests and unilateral/HA in the listening span and matrix test in quiet tasks. However, hearing condition only significantly predicted scores on listening span and matrix test in quiet. Previous literature has investigated the spatial hearing effects of using one CI versus two and this study corroborates their findings where bilateral users receive benefits in speech perception due to better sound localization (Best et al., 2015; Dunn et al., 2010; Glyde et al., 2013; Loizou et al., 2009; Perreau et al., 2017; A. Schäfer et al., 2011; Smulders et al., 2016; Tyler et al., 2007; van Hoesel, 2015; Wackym et al., 2007).

These studies suggest that the benefit might stem from having the ability to segregate and attend to a target speaker or sound stimuli which can explain why bilateral users, in this study and past studies, have reported better enjoyment and clarity of media (Dorman et al., 2008; Gfeller et al., 2008; Kong et al., 2005; Veekmans et al., 2009), lower listening effort (Dunn et al., 2010; Hughes & Galvin, 2013; Noble et al., 2008; Perreau et al., 2017; Schnabl et al., 2015; Sladen et al., 2018), and better speech, spatial and quality of hearing (Kocak Erdem & Ciprut, 2019; Noble et al., 2008; Summerfield et al., 2006). Lower levels of listening effort and better quality of hearing suggests that the bilateral CI use allows for less cognitive resources to be used for encoding stimuli perhaps due to its spatial hearing advantage (Hua et al., 2017; Noble et al., 2008; Schnabl et al., 2015). This may be the reason why bilateral users also showed greater working memory ability, since more resources were available for other processes such as the understanding of speech for speech perception.

Surprisingly, clinical AzBio speech perception tests, did not find this difference between conditions; previous studies investigating speech perception in noise when speech and noise were presented from the same speaker found similar results (Laszig et al., 2004; Litovsky et al., 2006, 2009) however, once stimuli were separated, bilateral users performed better than the unilateral only group (Dunn et al., 2010). The reason for the discrepancy between the clinical speech perception tests and the online matrix tests is unclear where, the trend is similar showing bilateral users performing better than the other groups, the results however, are not significant with the clinical scores. Perhaps because the clinical speech perception tests are more difficult compared to the matrix tests. AzBio sentences are longer and there exists more variability in performance within each hearing condition and therefore, the differences between hearing conditions become less robust. Although these differences exist between hearing conditions and contribute to predicting performances on online working memory and speech perception tests, they do not seem to predict survey and clinical speech perception outcomes; what we find is that working memory, speech perception and certain demographic variables, other than hearing condition, are more predictive.

### 4.3. Working memory and speech perception related to clinical and survey outcomes

#### 4.3.1. Speech perception and quality of hearing

This study demonstrated that the listening span in noise test and duration of deafness in CI users were able to predict AzBio in noise scores where the working memory test was positively correlated, and duration of deafness was negatively correlated. Also, that the online working memory and speech perception tests were able to explain variability in subjective speech hearing and quality of hearing outcomes where all were positively correlated. This suggests that a stronger working memory ability and a lower duration of deafness, lead to better scores on the speech perception and better working memory and speech perception leads to higher reports of ease of listening, clarity and identifiability of different speech and sound stimuli.

In a social setting, hearing-impaired individuals before and after obtaining a CI, rely on other cues such as lip reading, gestures, and other visual speech cues to understand speech (Bishop & Miller, 2009; Rouger et al., 2008; Song et al., 2015). These cues require more cognitive resources to interpret and use them to disambiguate the speech stimuli (Hirst et al., 2018; Mattys et al., 2012; Ohlenforst et al., 2017; Rönnberg et al., 2010, 2016; Schorr et al., 2005; Stropahl et al., 2017; Sumby & Pollack, 1954). In consequence, the longer they had profound deafness before obtaining a CI, the more changes that may occur neurologically in terms of sensory processing and cognition. In terms of sensory processing, studies have found smaller auditory cortical activity in response to auditory stimuli with increasing length of duration of deafness (Green et al., 2005; Han et al., 2019; Ito, 1993; D. S. Lee et al., 2001; H.-J. Lee et al., 2007; Sun et al., 2021). Along with that, studies have shown increased activation in the auditory cortex in response to visual stimuli where, in other studies, it was negatively correlated with speech perception and not duration of deafness (Anderson et al., 2019; Buckley & Tobey, 2011; Chen et al., 2016; Doucet et al., 2006; Giraud & Lee, 2007; Sandmann et al., 2012; Schierholz et al., 2015; Strelnikov et al., 2013). This cross-modal plasticity can occur due the diminished input of signals into the auditory cortex in combination with the increased reliance on visual cues resulting in CI users processing visual input using their auditory cortex (Buckley & Tobey, 2011; Chen et al., 2016; Doucet et al., 2006; Finney et al., 2003; Sandmann et al., 2012; Strelnikov et al., 2013; Stropahl et al., 2015).

Studies have also shown the increased activation of frontal regions, specifically the inferior frontal gyrus, in speech perception, where larger activity is correlated with better performance (Eisner et al., 2010; Giraud et al., 2011; Kessler et al., 2020; Lee et al., 2007; Mortensen et al., 2006; Strelnikov et al., 2015; Suh et al., 2015). This change occurs before obtaining a CI and its level of activation before implantation is predictive of performance outcomes afterwards (Suh et al., 2015). These results may suggest that as a result of changes in sensory processing favouring visual stimuli, compensatory neurocogntive changes may occur in order to comprehend speech stimuli. Plasticity in the cognitive areas of the brain are different between individuals; possibly due to differences in cognitive and working memory capacities. Individuals with lower speech perception scores have been shown, in the present and other studies, to have lower working memory capacities (Moberly, Houston, et al., 2017). CI users with larger working memory capacities may have the ability to recruit more resources for selective attention (Coez et al., 2014; Kessler et al., 2020; Mertens et al., 2020) and for the comprehension and storage of speech stimuli (Eisner et al., 2010; Giraud et al., 2011; Kessler et al., 2020; Lazard et al., 2010b; H.-J. Lee et al., 2007; Moberly, Houston, et al., 2017; Mortensen et al., 2006; K. Strelnikov et al., 2015; Suh et al., 2015) than those with lower. These individuals with lower capacities may allocate more resources for the encoding and perception of speech stimuli however, since resources for working memory are limited, less becomes available for the understanding and storage of speech stimuli (Ala et al., 2020; Arlinger et al., 2009; Bashivan et al., 2014; F. R. Lin, Metter, et al., 2011; Marsella et al., 2017; Miles et al., 2017; Pichora-Fuller et al., 2016; Pichora-Fuller & Singh, 2006; Rudner et al., 2011; Scheeringa et al., 2009; Xie et al., 2016).

#### 4.3.2. Spatial Hearing

The results of this study also show that working memory and speech perception online test scores, contribute to the variability found in subjective reports of spatial hearing for CI users and for NH individuals, online working memory and age contributed to the variability. Correlations suggested that for CI users, those with better working memory and speech perception, report better spatial hearing and NH individuals that are younger and with better working memory score, report better spatial hearing. Literature comparing spatial hearing ability between CI users and NH participants have only compared NH controls with bilateral CI users and showed a superior ability and better reports of spatial hearing (Kan & Litovsky, 2015). A theory to explain why sound localization is difficult for CI users is that to judge the location of a sound, NH individuals use interaural level differences (ILD) combined with interaural time differences (ITD), however, studies have shown CI users to rely more on the former than the latter (Aronoff et al., 2010, 2012; Grantham et al., 2007; Seeber & Fastl, 2008; van Hoesel & Tyler, 2003). In a study paradigm where participants simply had to indicate the direction of a sound, authors found that NH individuals and CI users, who performed better on the task, had a greater activation of their right temporo-parietal-occipital junction which is involved in orienting attention towards new stimuli (Schäfer et al., 2021). Other studies have shown that with intensive training using similar paradigms, spatial hearing can be improved (Firszta et al., 2014; Majdak et al., 2013; Yu et al., 2018).

Poor ability to localize a target sound with distractors present can result in an increased use of working memory resources for speech perception. When having to segregate streams of speech without ILD (volumes of competing speech streams were the same), bilateral CI users show difficulties in segregating streams of speech when spatially distinct which might be a reason why speech in noise is difficult for them. Neural correlates of this phenomenon are found in Paul et al., 2020; bilateral CI users and NH controls were told to attend to a left or right speaker and then, in both speakers, a set of digits were played, simultaneously or shifted in time, which they were asked to recall the digits (Paul et al., 2020). NH individuals, in this study, show a stronger early cortical entrainment in the superior temporal gyrus to an attended talker versus ignored whereas bilateral CI users show a weaker differentiation between the two talkers and a larger alpha power activity in the parietal cortex, ipsilateral to the distracting speaker. Alpha power indicates suppression of a neural areas to protect task-relevant stimuli by inhibiting the encoding of distracting stimuli (Bashivan et al., 2014; Bonnefond & Jensen, 2012; Jensen, 2002; Klimesch et al., 2007; Roux & Uhlhaas, 2014; Scheeringa et al., 2009; Tuladhar et al., 2007; Xie et al., 2016). In this case, the parietal cortex, which is involved in selective spatial attention, was suppressed more so in CI users than NH. This increased use of resources to suppress distracting stimuli shows the increased effort CI users use to segregate streams of speech even when they are spatially distinct, and it affects their ability to recall the presented target stimuli.

#### 4.3.3. CIQOL: Entertainment

Lastly, subjective reports of entertainment were predictable using performance on the working memory and speech perception tests. The entertainment section measured enjoyment and clarity of TV, radio and music and correlation analysis showed that variables positively correlated with the survey scores suggesting that CI users with better working memory and speech perception were able to enjoy media more. The perception of speech stimuli through TV and radio are not well documented compared to music perception in CI users. However, it has been seen clinically that CI users have difficulty understanding speech on TV and reasons for this are that people on TV may not be facing them or are off-screen, they speak too quickly, background noise or music, and distance listening (Clark, n.d.). Its relationship with individual differences in cognition and working memory, therefore, can only be speculative. In the case of this study, it suggests that those with a stronger and more efficient working memory and speech perception ability, find that media through TV and radio is clearer and more enjoyable than those with weaker.

Music perception has been shown to be highly variable in CI users (Drennan et al., 2015; Gfeller et al., 2008; Philips et al., 2012; Wright & Uchanski, 2012) and correlated with speech perception in noise where individuals with better speech perception in noise report higher levels of listening and enjoyment of music (Dincer D’Alessandro et al., 2021). Previous studies have found difficulties when investigating CI users listening to lyrical music; this may suggest that the instrument accompaniment of lyrical music might act as background noise while they attempt to hear and understand the lyrics (Collister & Huron, 2008; Eskridge et al., 2012; Gfeller et al., 2008, 2019). However, other studies have shown that perception of pitch and timbre in music are difficult for CI users; both of which are also elements of speech and promote pleasure and other emotions while listening to music (Limb & Rubinstein, 2012; McDermott, 2004; Moran et al., 2016) which might be why those with better speech perception also find more pleasure with music.

CI users that undergo music therapy have shown a positive effect in speech perception, music perception and appreciation of music (Firestone et al., 2020; Hutter et al., 2015; Looi et al., 2012). The reason for these results could be that brain regions involved in the perception of musical stimuli are shown to overlap with that of perception of auditory stimuli, attention and working memory (Looi et al., 2012; Patel, 2014; Shahin, 2011) therefore, when there are difficulties with one of the above, the others might have issues as well. Music therapy seems to allow for neural networks related to those processes to be more activated allowing for better speech perception in noise ability through enhanced auditory encoding, top-down processing, and cross-modal integration (Moran et al., 2016; Parbery-Clark et al., 2009).

### 4.4. Implications and future directions

This study showed the effects of hearing condition on working memory, speech perception and subjective survey outcomes and the effects of working memory, speech perception and demographics on subjective survey and clinical speech perception outcomes. The results suggest that those with better working memory and speech perception report better quality of life and hearing and those with better online test performance scores, tend to be bilateral CI users. It has been alluded to in past studies that better spatial hearing reduces the amount of cognitive effort used in a social setting due to a more efficient method of segregating and attending to a target speaker amidst background noise or distractors (Best et al., 2015; Dunn et al., 2010; Glyde et al., 2013; Loizou et al., 2009; Perreau et al., 2017; A. Schäfer et al., 2011; Smulders et al., 2016; Tyler et al., 2007; van Hoesel, 2015; Wackym et al., 2007). This suggests that the spatial advantage that a second CI provides, allows for an increase in working memory ability and a better ability to perceive speech. Although it is beneficial to have a second CI, their spatial and speech perception performance compared to NH individuals is still weaker and they use more cognitive resources due to differences in encoding spatially distinct information (Aronoff et al., 2010, 2012; Grantham et al., 2007; Kan & Litovsky, 2015; Paul et al., 2020; Seeber & Fastl, 2008; van Hoesel & Tyler, 2003) Future longitudinal studies on the cognitive effects of using bilateral versus unilateral CIs and compared to NH individuals are required to investigate the possible cognitive consequences of not obtaining a second CI and to see if bilateral CI users still present a pattern of cognitive decline and dementia later in life so commonly found in individuals with hearing loss (Lin, Metter, et al., 2011; Lin, Thorpe, et al., 2011; Livingston et al., 2017; Lopes et al., 2007; Slade et al., 2020). If that is that case then interventions involving the improvement of working memory and cognitive abilities in CI users, would possibly result in better outcomes and protection against cognitive decline.

### 4.5. Limitations

A main limitation in this study is the lack of control over stimuli presentation and environment. Since this was an online study, participants completed tasks using their own devices and in an environment of their choosing. The differences in speaker or headphone quality were not controlled for and while some CI users used speakers, others used a Bluetooth or wired connection to stream the audio directly to their processor which may provide added benefits compared to listening through speakers. In terms of environment, participants did not complete the studies in a soundproof booth and therefore, we cannot confirm that they completed the study without any external distractions.

### 4.6. Conclusion

We investigated the effects demographics on the performance of online working memory and speech perception tests along with the effects of online tests and demographics on quality of life, quality of hearing and clinical speech perception outcomes. Our findings support previous literature suggesting that the use of bilateral CIs not only present benefits to speech perception but also to working memory ability, enjoyment and clarity of media, listening effort, speech, spatial and quality of hearing where they were superior to unilateral/HA and unilateral only users (Dorman et al., 2008; Dunn et al., 2010; Gfeller et al., 2008; Hughes & Galvin, 2013; Kocak Erdem & Ciprut, 2019; Kong et al., 2005; Noble et al., 2008; Perreau et al., 2017; Schnabl et al., 2015; Sladen et al., 2018; Summerfield et al., 2006; Veekmans et al., 2009) We also find that CI users with better working memory ability and shorter durations of deafness, predicted better speech perception in noise. In addition to that, those with better working memory and speech perception, report better speech hearing, spatial hearing, quality of hearing and enjoyment and clarity of media. Overall, these findings show that the improvement of working memory and speech perception in CI users is important in ensuring better speech perception, quality of life and quality of hearing. The detriments in working memory and speech perception are affected by the type of hearing condition where the use of a second CI and therefore, its added spatial hearing advantage, allows for a reduction on the cognitive burdens of listening and an increase in performance on both. Therefore, a second CI along with training and improvement in working memory may provide protection from cognitive decline later in life.

## Data Availability

All data produced in the present study are available upon reasonable request to the authors

## Acknowledgements

The authors would like to thank Emmanuel Chan and Lina Khayyat for their assistance with participant recruitment, data collection, and compensation.

